# Blood Pressure Variability and Autonomic Response to an Acute Bout of High Intensity Interval Exercise in Healthy Young Adults

**DOI:** 10.1101/2024.01.29.24301957

**Authors:** Saniya Waghmare, Alicen A. Whitaker-Hilbig, Mark Chertoff, Sandra A. Billinger

## Abstract

Autonomic nervous system (ANS) activity causes acute variations in the blood pressure. Blood pressure responds to high intensity interval exercise (HIIE) repeatedly during alternating intensities, however, ANS response to the changing intensities of HIIE is unknown. We characterized the response of beat-to-beat blood pressure variability (BTB BPV) to an acute bout of HIIE using coefficient of variation (CoV) and spectral low frequency [LF], and high frequency [HF] domains. Our hypotheses were mean arterial pressure BTB BPV, would increase during 1) high intensity and 2) active recovery of HIIE compared to baseline (BL). BTB BPV would reduce during 1) cool down 2) post HIIE 3) 30 minutes post HIIE compared to BL in young adults. HIIE included bouts of 1-minute high-intensity separated by 1-minute recovery (□70% and 10% estimated Wattmax) for total of 10 minutes on a recumbent stepper. A secondary analysis was performed using twenty-one datasets of young individuals (age 25±1.5, 48% female). During high intensity, LF and HF increased compared to BL (p < 0.05) indicating increased sympathetic activity and breathing. During active recovery, LF and HF remained elevated above BL and were greater than during high intensity (p ≤ 0.02). Sympathetic activity reduced back to BL immediately post HIIE but returned to being higher than BL at 30 minutes after HIIE (p=0.001). BTB BPV CoV also increased during HIIE compared to BL (p<0.05). Results suggest that young healthy individuals have increased BTB BPV during HIIE suggesting cardiovascular system responds to ANS fluctuations during changing exercise intensity.

**New and Noteworthy:** This novel study analyzed beat -to-beat blood pressure variability during high intensity interval exercise (HIIE) in young healthy adults. We found that blood pressure variability was highest during active recovery compared to resting or high intensity exercise. Moreover, variability increased during HIIE but returned to resting post-exercise. These findings provide valuable insights into the blood pressure and ANS responses to HIIE, contributing to our understanding of their impact on overall cardiovascular health in young adults.

## INTRODUCTION

The autonomic nervous system (ANS) through the sympathetic and parasympathetic branches regulates key involuntary physiologic processes such as heart rate, blood pressure, and breathing at rest and during physiologic challenges such as exercise.(1–3) During exercise, the ANS regulates blood pressure by increasing sympathetic nervous activity within the vasculature to maintain a blood pressure gradient while increasing blood flow to provide oxygen and nutrients to the muscles and vital organs.(2, 4, 5) The physiological changes in blood pressure due to the ANS activity can be assessed using beat-to-beat blood pressure variability (BTB BPV).(5–7) BTB BPV has been previously characterized during continuous incremental exercise from low to high intensity.(5, 8) During increases in exercise intensity, sympathetic nervous activity and breathing frequency increases, which in turn increases BTB BPV.(5, 8) However, it is unknown how interval exercise may influence the ANS and BTB BPV in young healthy adults.

High intensity interval exercise (HIIE) includes alternating between high intensity exercise and recovery bouts (9, 10) and has been shown to influence the cardiovascular system greater than moderate intensity exercise.(10–12) While blood pressure has been shown to change concomitantly with exercise intensity during short-interval, low-volume HIIE in young healthy adults,(13) blood pressure alone does not provide sufficient evidence about the ANS function and cardiovascular hemodynamics. Characterizing BTB BPV during short-interval, low-volume HIIE in young healthy adults would provide unique insight about the ANS and its influence on the cardiovascular system during rapid changes in exercise intensity.(6, 7, 14) As sympathetic activity is intensity dependent, one would anticipate increases in sympathetic activity during HIIE which will increase BTB BPV to maintain perfusion pressure. During the recovery bouts of HIIE, decreases in exercise intensity and sympathetic activity may therefore reduce BTB BPV.(15, 16) However, BTB BPV may not return to resting levels during the active recovery bouts due to the continued exercise at low-intensity which would maintain increased sympathetic activity for the short 1-min duration. However, there wouldn’t be a withdrawal of sympathetic activity and a rapid decrease in blood pressure until the cool-down phase immediately following HIIE when both exercise intensity and cadence are decreasing back to a resting state.(13) To reduce the occurrence of post-exercise hypotension and syncope, the ANS may quickly return back to the resting sympathovagal balance following exercise. (17, 18) Therefore, understanding autonomic function and BTB BPV during changing intensities of HIIE and recovery in healthy young adults would provide insights for cardiovascular hemodynamic responses and provide a framework for comparison to people living with cardiovascular or neurological disease.

The primary aim of this investigation was to examine changes in BTB BPV during and following a single bout of HIIE in young, healthy adults. Our primary hypothesis suggests that BTB BPV parameters such as the co-efficient of variation (CoV), low frequency (LF) power spectral density (PSD), high frequency (HF) PSD, and the LF/HF ratio will increase during both the high-intensity phase of HIIE, and its active recovery phase compared to baseline (BL). The secondary hypothesis anticipates reductions in BTB BPV CoV, LF PSD, HF PSD, and the LF/HF ratio during the cool down following HIIE, at rest immediately following HIIE, and 30 minutes after HIIE, relative to BL.

## METHODS

We performed a secondary analysis of existing data where the primary focus was cerebrovascular response to HIIE.(13) Individuals enrolled into the study provided written informed consent prior to the study visits and were: 1) age 18–30 years old and 2) low cardiac risk defined by the American College of Sports Medicine (ACSM).(19) For those included in this secondary analysis, participant characteristics are described in Table 1. The primary study was approved by University of Kansas Medical Center Human Subjects Committee.

**Table 1.** Participant Characteristics.

|  | **All** | **Women** | **Men** | **p value** |
| --- | --- | --- | --- | --- |
| **n** | 21 | 10 | 11 |  |
| **Age (years)** <sup>∞</sup> | 24.7 (SD 1.5) | 24.6 (SD 1.9) | 24.9 (SD 1.0) | 0.65 |
| **BMI, Kg/m<sup>2</sup>** <sup>∞</sup> | 23.6 (SD 3.6) | 21.0 (3.26) | 26.0 (1.89) | <0.001* |
| **Race/ethnicity** <sup>□</sup>, n (%) |  |  |  |  |
| **White/ Caucasian** | 17 (81.0%) | 9 (90.0%) | 8 (72.7%) | 1.00 |
| **Hispanic/Latino** | 1 (4.8%) | 0 | 1 (9.1%) | 1.00 |
| **Asian** | 3 (14.3%) | 1(10%) | 2 (18.2%) | 1.00 |
| **Native American** | 1(4.8%) | 0 | 1 (9.1%) | 1.00 |
| **Physical Activity score** <sup>□</sup>, n (%) |  |  |  |  |
| **<10 min, 5 time/wk** | 1(4.8%) | 1(4.8%) | 0 |  |
| **20-60 min/wk** | 1 (4.8%) | 0 | 1 (9.1%) |  |
| **1-3 h/wk** | 1 (4.8%) | 0 | 1 (9.1%) |  |
| **>3 h/wk** | 18 (85.7%) | 9 (90%) | 9 (81.8%) | 1.00 |
| **Estimated VO<sub>2max</sub>** <sup>∞</sup>, ml.kg<sup>-1</sup>.min<sup>-1</sup> | 44.6 (SD 6.4) | 41.9 (SD 4.5) | 47.1 (SD 7.1) | 0.06 |

Means (standard deviation). \*Significant between groups (p<0.05). ml.kg<sup>-1</sup>.min<sup>-1</sup> = milliliters of oxygen per kilogram of body weight per minute. VO<sub>2max</sub> = maximum oxygen consumption. BMI = Body mass index. Kg/m<sup>2</sup> = kilogram of body weight per meter square. h/wk = hours per week. min/wk = minute per week. <sup>□</sup> Analyzed by Fisher exact test. <sup>∞</sup>Analyzed by one way ANOVA.

The methodology for the primary paper are presented in greater detail elsewhere. (13) Briefly, participants performed a submaximal exercise test using a recumbent stepper (20, 21) to determine the workload for the HIIE bout during their visit 1.(13, 22) Following a 20-minute rest on the recumbent stepper, participants were instrumented with equipment. For purposes of this study, we present the following variables of interest: 1) 5-lead electrocardiogram (ECG; Cardiocard, Nasiff Associates, Central Square, NY) for HR, and 2) beat-to-beat BP finger cuff (Finometer, Finapres Medical System, Amsterdam, The Netherlands) for mean arterial pressure (MAP).

Data was continuously recorded for 5 minutes of seated rest at baseline (BL). The HIIE protocol started with active recovery to decrease the effect of a Valsalva maneuver.(23) The HIIE bout consisted of repetitive 1-min high intensity (70% estimatedWattmax) separated by 1-min active recovery (10% estimatedWattmax).(13) The short-interval, low-volume HIIE bout alternated every minute between the high-intensity at 70% Wattmax and the active recovery at 10%Wattmax. After 10 minutes of HIIE, resistance was decreased to 15 W for the 2 minutes of cooldown. Five-minute recordings commenced in a seated rest position at 5- and 30-minutes post-HIIE.

## AUTONOMIC RESPONSE AND VARIABILITY ASSESSMENT

The data was collected using an analog-to-digit unit (NI-USB-6212, National Instruments) and custom written MATLAB code (v2017a, The MathWorks, Inc., Natick, MA) using methods described previously.(13, 22) Raw data were sampled at 500Hz and then resampled at 10Hz. We extracted the data for beat-to-beat mean arterial pressure (MAP) (Finometer, Finapres Medical System, Amsterdam, The Netherlands). We obtained power spectral density by using custom written MATAB code for low and high frequencies. To examine the influence of the autonomic nervous system on BP, we measured BTB BPV in the temporal domain using beat-to-beat mean arterial pressure (MAP) CoV and in the spectral domain using LF PSD and HF PSD. PSD of BTB BPV obtained across the low frequency band (0.04Hz to 0.15Hz) is influenced by sympathetic nervous activity and vascular modulation. PSD of BTB BPV within the high frequency band (0.15Hz to 0.40Hz) is influenced by breathing or parasympathetic activity.(14, 24, 25) The ratio of low and high frequency (LF/HF ratio) measures the balance between sympathetic and parasympathetic nervous activity, also known as sympathovagal balance.(24–26)

The power spectral density was determined using Fast Fourier Transform (FFT) analysis applying cross power spectral density (cPSD) software package, 100-s Hanning windows, and 50% superposition. The power spectral density algorithm of MATLAB converted the time domain of MAP to frequency. We detrended the data and used cPSD to obtain PSD during HIIE. For appropriate resolution for PSD during HIIE, we analysed PSD of high-intensity bouts of HIIE by concatenating resampled data of every high intensity bout minute to obtain 5-minutes of data shown in **Supplementary Figure 1**. Similarly, we analyzed the combined active recovery bouts of HIIE as 5-minutes of resampled data. Aligned with previous studies, the peak power of MAP was determined across the power spectrum from 0.04Hz to 0.15Hz for LF and 0.15Hz to 0.40Hz for HF.(14, 26) The changes of power spectral density of MAP across time point are shown in **Supplementary Figure 2**. We then calculated the ratio of LF/HF. We calculated absolute mm^2^Hg/Hz and area under the curve (AUC) mm^2^Hg/Hz. For absolute BTB BPV we took the sum of PSD values within the LF and HF bands. AUC was calculated by the trapezoidal method similar to mentioned in other studies.(27, 28) We analysed the area by the height and width of the LF and HF peaks across PSD. For the time domain analysis, we calculated BTB MAP CoV using the equation:(29)

CoV= (Standard deviation/ Mean) * 100

## DATA ANALYSIS

Data analysis was performed using statistical package of social sciences (SPSS 27 version for Windows: SPSS Inc. Chicago, IL). A priori α was set as < 0.05. We assessed the normality and sphericity using Shapiro Wilk test and Mauchly’s test, respectively. Participant’s characteristics were analysed using one-way ANOVA, and Fisher’s exact test for categorical variables. BTB BPV CoV were analyzed across time using repeated measures ANOVA for normally distributed data, and Friedman’s test for non-normally distributed spectral measures of LF, HF, LF:HF ratio (AUC and absolute). Post hoc comparisons used paired t-tests and Wilcoxon’s signed rank tests.

## RESULTS

Twenty-five datasets were available for our secondary analysis. We excluded 4 datasets due to noise in signal and therefore had a sample of 21 complete data sets. Since, this was a secondary analysis, an apriori sample size was not calculated. Other studies that reported blood pressure variability during an acute bout of continuous incremental exercise reported a sample size of 15 participants which is smaller than the sample in this analysis.(5, 8) .

### POWER SPECTRAL DENSITY (PSD)

All spectral measures of BTB BPV significantly changed across time, as shown in Table 2.

**Table 2.**
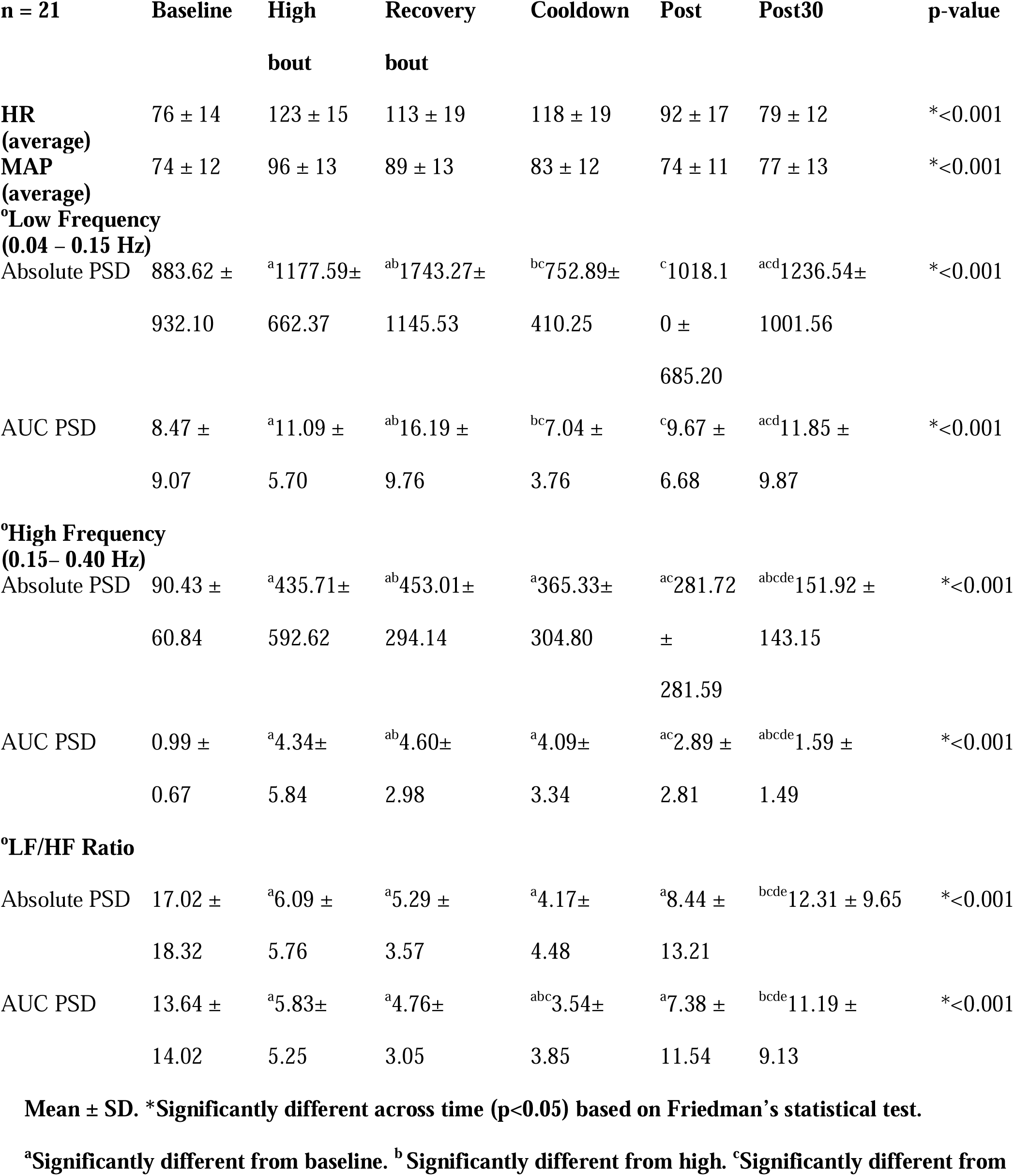
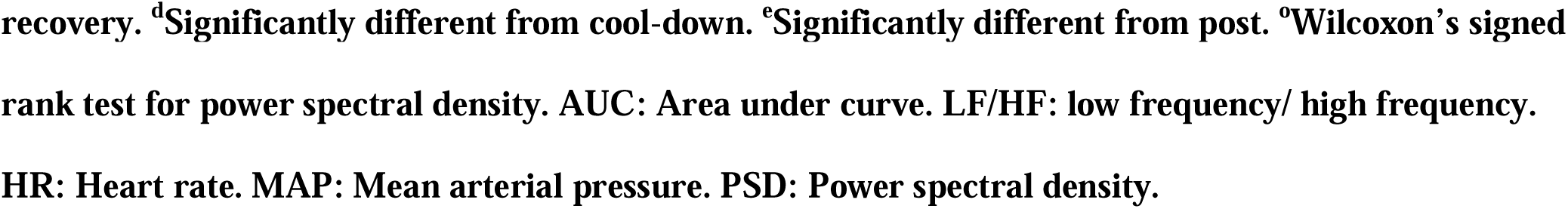
Power spectral density of blood pressure variability across timepoints.

#### LOW FREQUENCY (LF)

LF AUC PSD significantly increased during the high-intensity bouts of HIIE compared to BL (p = 0.016) and stayed higher during active recovery (p < 0.001) as shown in **Figure 1**. During high intensity of HIIE, LF AUC PSD was lower than active recovery (p < 0.01). During cooldown and immediately post HIIE, LF AUC PSD returned to BL and was not significant (p= 0.972, p = 0.122). At 30-minutes after HIIE, LF AUC PSD once again increased above BL (p = 0.002).

**Figure 1.**
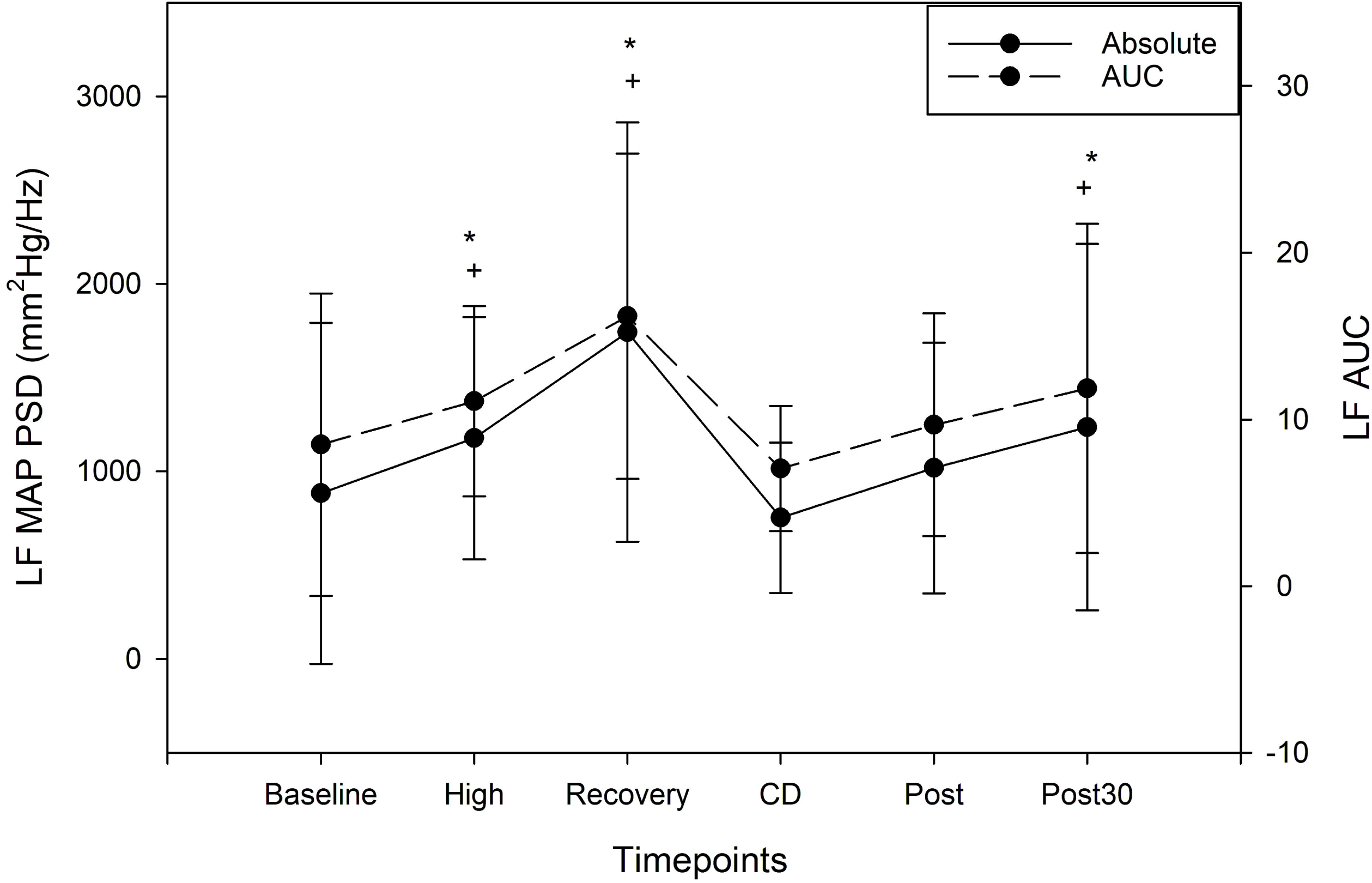
Beat-to-beat blood pressure variability (MAP) power spectral density (Low Frequency) response to an acute bout of high intensity interval exercise (HIIE, n = 21); *Significantly different than baseline (rest) absolute power spectral density (p < 0.005). ^+^Significantly different than baseline (rest) area under curve of power spectral density (p < 0.005). MAP, mean arterial pressure; LF MAP PSD, low frequency mean arterial pressure power spectral density absolute values (closed dots); LF AUC, low frequency area under curve (open dots); Baseline, rest; High, low frequency during high intensity of HIIE; recovery, low frequency during active recoveries of HIIE; CD, active cool down; Post, immediately post HIIE; Post 30, post 30 minutes after HIIE; mm^2^Hg/ Hz millimetre square of mercury per hertz.

LF absolute PSD followed the same pattern as LF AUC PSD. LF absolute PSD significantly increased during the high intensity bouts of HIIE compared to BL (p= 0.03) and stayed higher during active recovery (p<0.001) as shown in **Figure 1**. Compared to active recovery, LF absolute PSD during high intensity of HIIE was lower (p < 0.001). During cool down and immediately post HIIE, LF absolute PSD returned to near BL and was not significant (p= 0.76, p = 0.14). At 30-minutes after HIIE, LF absolute PSD once again increased from BL (p < 0.001).

#### HIGH FREQUENCY (HF)

HF AUC PSD was significantly increased during the high-intensity bouts of HIIE compared to BL (p < 0.001) and stayed higher during active recovery (p < 0.001) as shown in **Figure 2**. HF AUC PSD during high intensity of HIIE was lower compared to active recovery (p = 0.02). During cooldown and immediately post HIIE, HF AUC PSD stayed higher than BL (p < 0.001). At 30 minutes after HIIE, HF AUC PSD remained increased above BL (p= 0.012).

**Figure 2.**
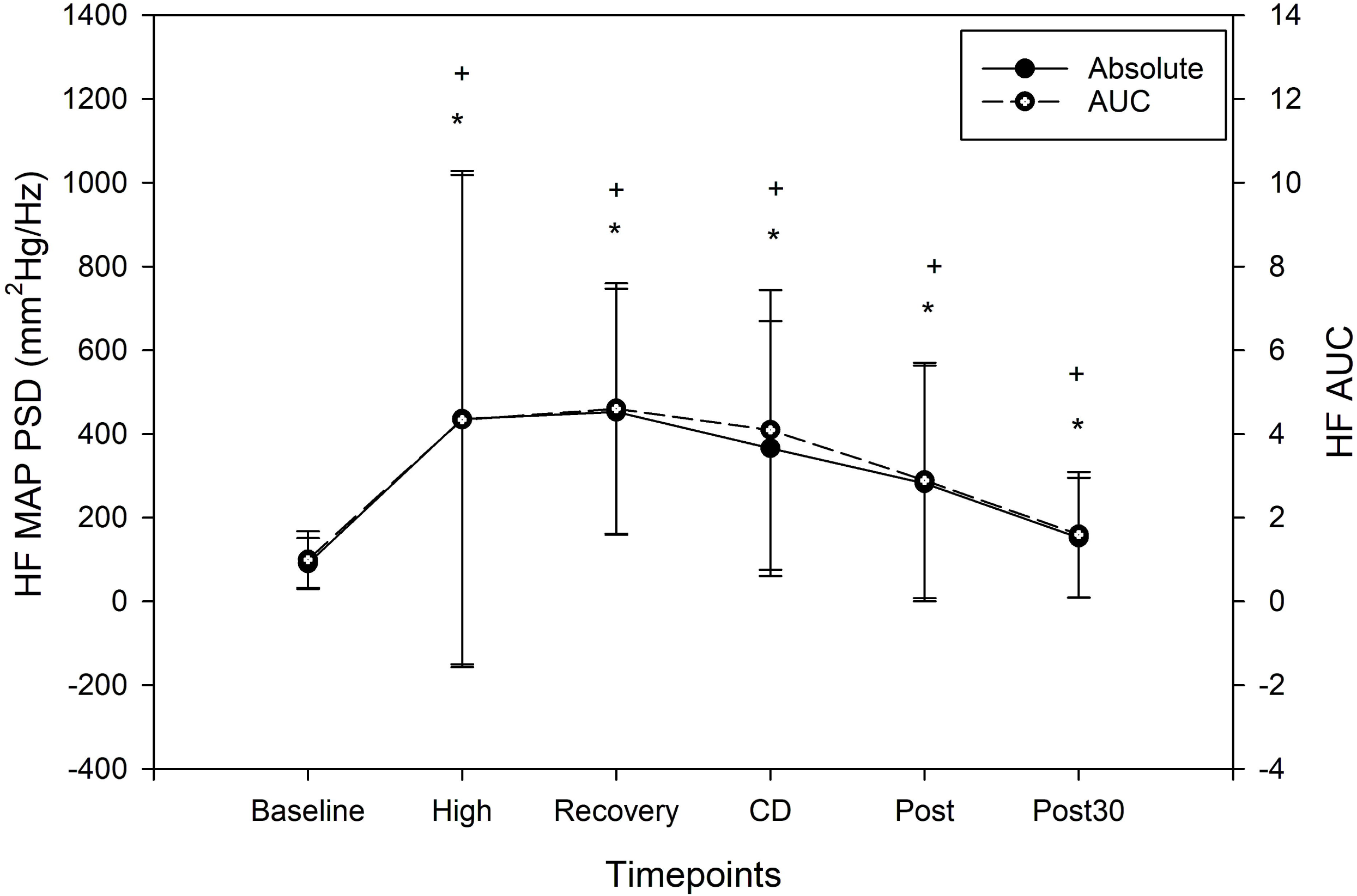
Beat-to-beat blood pressure variability (MAP) power spectral density (High Frequency) response to an acute bout of high intensity interval exercise (HIIE, n = 21); *Significantly different than baseline (rest) absolute power spectral density (p < 0.005). ^+^Significantly different than baseline (rest) area under curve of power spectral density (p <0.005). MAP, mean arterial pressure; HF MAP PSD, high frequency area under curve (open dots); Baseline, rest; High, high frequency during high intensity of HIIE; recovery, high frequency during active recoveries of HIIE; CD, active cool down; Post, immediately post HIIE; Post 30, post 30 minutes after HIIE; mm^2^Hg/ Hz millimetre square of mercury per hertz.

Once again, HF absolute PSD followed the same pattern as HF AUC PSD. HF absolute PSD was significantly increased during high-intensity bouts of HIIE (p <0.001) and stayed higher during active recovery bouts of HIIE (p < 0.001). Compared to active recovery, HF absolute PSD during high intensity of HIIE was lower (p = 0.025). During cooldown and immediately post HIIE, HF absolute PSD stayed higher than BL (p < 0.001), (p < 0.001). At 30 minutes after HIIE, HF absolute PSD stayed increased from BL (p=0.011), see **Figure 2**.

#### LF/HF RATIO

The LF/HF ratio AUC significantly reduced during high intensity bouts of HIIE (p=0.007) compared to BL and stayed reduced during active recovery bouts (p=0.001) as shown in **Figure 3**. There was no significant difference between active recovery and high intensity of HIIE (p = 0.27). During cool down and immediately post HIIE, LF/HF ratio AUC remained lower than BL (p=0.001, p=0.013, respectively). However, LF/HF ratio AUC returned to BL values at 30 minutes after HIIE (p=0.84).

**Figure 3.**
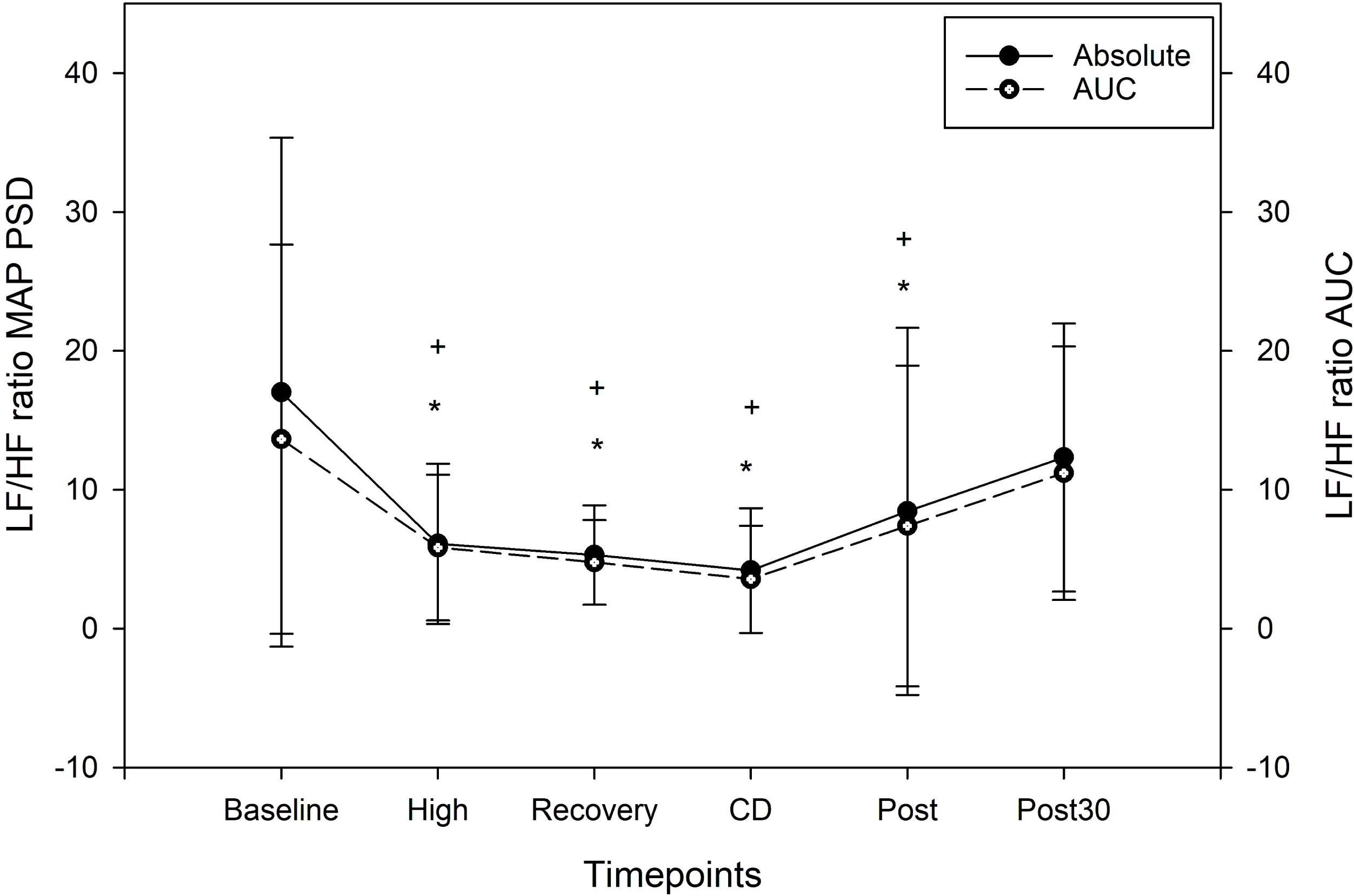
Beat-to-beat blood pressure variability (MAP) power spectral density (LF/HF ratio) response to an acute bout of high intensity interval exercise (HIIE, n = 21); *Significantly different than baseline (rest) absolute power spectral density (p<0.005). ^+^Significantly different than baseline (rest) area under curve of power spectral density (p<0.005). MAP, mean arterial pressure; ratio MAP PSD, low and high frequency ratio area under curve (open dots); Baseline, rest; High, ratio of frequencies during high intensity of HIIE; Recovery, ratio of frequencies during active recoveries of HIIE; CD, active cool down; Post, immediately post HIIE; Post 30, post 30 minutes after HIIE; mm^2^Hg/ Hz millimetre square.

Once again, the pattern of LF/HF ratio absolute PSD was the same as the AUC. The LF/HF ratio absolute significantly reduced during the high-intensity bout of HIIE (p=0.001) and stayed reduced during active recovery of HIIE (p=0.001) compared to BL. During cool down and immediately post HIIE, LF/HF ratio absolute PSD remained lower than BL (p=0.00, 1p=0.012, respectively). LF/HF ratio absolute PSD returned to BL post 30 minutes after HIIE (p=0.79) as shown in **Figure 3**.

#### CO-EFFICIENT OF VARIATION (CoV)

There was significant change in BTB BPV CoV across time (p < 0.001). Compared to BL, BTB BPV CoV was increased during HIIE (p < 0.05), except active recovery bout minute 1, high-intensity minute 4, and high-intensity minute 6 were not significantly different than BL. While BTB BPV CoV increased during HIIE, it did not follow a consistent pattern when switching between high-intensity and active recovery. **Figure** 4 shows the BTB BPV CoV returned to BL immediately following HIIE and 30 minutes after HIIE (p= 0.179 and p= 0.198 respectively).

**Figure 4.**
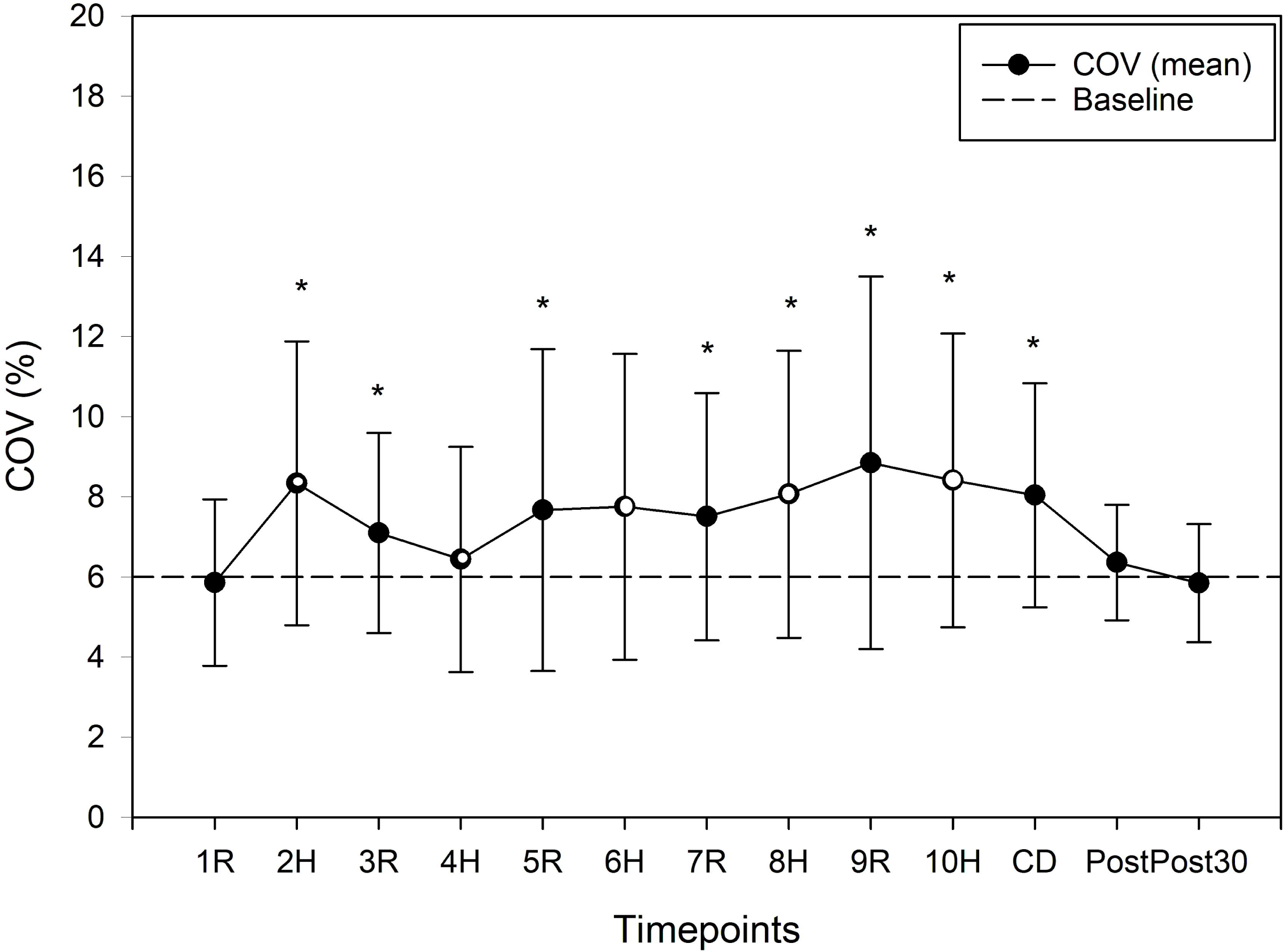
Coefficient of variation of beat-to-beat blood pressure variability (MAP) response to high intensity interval exercise (HIIE, n = 21); Horizontal dashed line represents baseline value. *Significantly different than baseline (p<0.005). MAP, mean arterial pressure; COV, coefficient of variation; H, high intensity (open dots); R, active recovery (closed dots); CD, active cool down; Post, immediately post HIIE; Post 30, post 30 minutes after HIIE.

## DISCUSSION

While BPV has been characterized during continuous exercise,(5, 8, 30) to our knowledge our study is the first to examine the acute effect of HIIE on BTB BPV and autonomic function. Our main findings were: 1) during the high intensity and active recovery of HIIE there was a significant increase in LF and HF MAP PSD compared to BL 2) and interestingly LF and HF PSD was lower during the high intensity bouts of HIIE compared to active recovery 3) in contrast to our hypothesis, during cool down and immediately post HIIE, LF PSD returned to BL while HF PSD remained elevated, and 4) at 30 minutes after HIIE, both LF and HF PSD remained higher than BL. Due to greater increase in HF PSD compared to LF PSD, the sympathovagal balance, measured as the LF/HF ratio, reduced below BL, during HIIE, cool down, immediately post HIIE and returned back to BL at 30 minutes after HIIE. A significant increase in BTB BPV CoV was seen across HIIE, with BTB BPV CoV returning to BL immediately post and 30 minutes after HIIE. Our findings of increased BTB BPV during HIIE are supported by previous studies showing increases in BTB BPV during continuous exercise.(8, 31)

### LF PSD during HIIE

The LF PSD increased during high intensity and active recovery of HIIE, indicating both sympathetic and vascular modulation increased BTB BPV.(32) Interestingly, the LF PSD during the high intensity of HIIE was lower than the active recoveries. Previous studies performing incremental low to high continuous exercise (5) report concomitant increases in BTB BPV with increasing intensity, but we did not show this during HIIE. One potential explanation for higher LF PSD during active recovery compared to high-intensity could be due to a latency effect or delay in sympathetic activity during exercise by appoximately 30-60 seconds.(1, 33, 34) The duration of active recovery bouts could have been too short to allow for a decrease in sympathetic activity. Another potential explanation to BTB BPV being lower during high-intensity could be due to the rapid increase in BP during high intensity causing baroreceptors to stretch, leading to a reduction in sympathetic activity within the vasculature. However, rapid decreases in BP during active recovery may have allowed the baroreceptors to once again increase the sympathetic activity within the vasculature and LF PSD to increase.(1, 35, 36) Further several metabolites released during high intensity such as lactic acid, potassium, arachidonic acid, and hydrogen ions could also contribute to increased sympathetic activity.(1, 37) The increased blood flow achieved by vasodilation washouts the metabolites to maintain pH, that may explain reduced LF PSD during high intensity.(8, 35–37) We acknowledge that factors that may also contribute to increased LF PSD during active recovery is renin-angiotensin system that releases norepinephrine at vasculature which in turn elevates sympathetic activity throughout heavy exercise.(8, 35–37)

### HF PSD during HIIE

HF PSD increased during high-intensity and active recovery of HIIE compared to BL. HF PSD is a measure of the parasympathetic activity and respiration’s influence on BTB BPV. HF PSD during rest measures vagal parasympathetic activity which mediates fluctuations of heart rate on BP causing variability.(38) During exercise parasympathetic activity withdraws and sympathetic activity increases which in turn increases heart rate and blood pressure.(39, 40) Therefore, increased HF PSD during HIIE may be a reflection of increased breathing rather than parasympathetic activity and requires further exploration.

### LF and HF during recovery

Our findings during cool down and immediately post HIIE determined LF PSD values returned to BL while HF PSD remained elevated. Previous studies have stated that during post exercise recovery the sympathetic activity withdraws, causing decreases in BP which may lead to post exercise hypotension.(41–43) Our results show sympathetic activity (LF PSD) returned to BL during the HIIE cool down and immediately post HIIE, however, it did not decrease below BL as hypothesized. Sympathetic activity increased 30 minutes after HIIE compared to BL, potentially to restore the sympathovagal balance and maintain tone of the blood vessel. HF PSD remained higher than BL immediately post HIIE and 30 minutes after HIIE. Post-exercise hyperapnea due to exercise oxygen debt leads to increased work of breathing and cardiac output, which may help explain the elevated HF PSD up to 30 minutes after HIIE.(44, 45)

### LF/HF Ratio (Sympathovagal balance)

In line with previous studies (42, 46) we did observe reduced sympathovagal balance during HIIE. However, there is a lack of consensus whether the LF/HF ratio is an accurate measure of sympathovagal balance during exercise.(8) Therefore, the LF/HF ratio may only be interpretable at BL and during recovery following HIIE but not during HIIE. As expected, we did observe reduced LF/HF ratio during cool down and immediately post HIIE. Further, LF/HF ratio at 30 minutes after HIIE returned to BL, to restore the sympathovagal balance.(42, 47) A quick return of sympathovagal balance back to BL could be a protective mechanism from further declines in blood pressure during post-exercise recovery which may induce syncope, as stated by a previous study examining LF/HF ratio increases during orthostasis.(48)

### Coefficient of variation during HIIE and recovery

BTB BPV CoV was also increased during HIIE. Interestingly, while MAP increased during high intensity and decreased during active recovery during HIIE,(13) we did not observe a similar pattern in the BTB BPV CoV. This finding may be due to the influence of many different physiological variables on BTB BPV CoV during exercise, including neurotransmitters released by the autonomic nervous system at blood vessels,(1, 49) changes of intrathoracic pressure,(3, 45) cardiovascular response based on the physical fitness,(50, 51), and baroreceptor sensitivity.(1, 52) Immediately post HIIE and 30 minutes after HIIE, BTB BPV CoV returned to BL, indicating a quick restoration of BP regulation following HIIE in young healthy adults.

## LIMITATIONS

To gain reliable results of BTB BPV over 5 minutes, we concatenated each minute of high intensity and active recovery bout and used the data for analysis, future studies can consider longer HIIE bouts to obtain ample data for analysis specifically for examining alternating intensity of high and recovery bouts of HIIE effects on BTB BPV. Due to the nature of secondary analysis, data was not available regarding baroreceptor sensitivity, which would provide information about baroreceptor role influencing blood pressure response during HIIE, this should be considered in future studies.

## FUTURE CONSIDERATIONS

Our results suggest that in young health individuals, HIIE as an exercise stimulus, increased autonomic function and the cardiovascular system was able to rapidly respond with greater BTB BPV. Future studies should examine mechanisms such as cardiac output, vasomotor activity, baroreflex sensitivity, and breathing contributions to the changes in BTB BPV during HIIE.(2, 8, 53, 54) Autonomic dysfunction in various other populations such as diabetes and heart failure may also respond to HIIE differently and provide clinical insights about the cardiovascular and autonomic function interaction.(55–57) Our analysis performed low volume HIIE, however future studies could examine effects of short and long interval HIIE which would provide different stimulus to factors that alter the BTB BPV response. (58)

## CONCLUSION

We show that HIIE increased autonomic modulation of blood pressure in terms of BTB BPV assessed by LF PSD, HF PSD, LF/HF ratio, and CoV. We also show BTB BPV was responsive to changes in intensity when switching between high intensity and active recovery bouts during HIIE in young healthy adults. BTB BPV and autonomic function returned to baseline values by 30-minutes after HIIE. Therefore, this study in young healthy individuals lays the foundation for characterizing a healthy BTB BPV and autonomic response to HIIE. Further research characterizing the BTB BPV and autonomic response to HIIE is needed in clinical populations such as heart disease and stroke due to underlying hypertension and reduced autonomic function.

## DATA AVAILABILITY

Data will be made available upon reasonable request.

## AUTHORS CONTRIBUTIONS

S.W., A.A.W.H., and S.A.B. conceived and designed research; A.A.W.H. performed experiments; S.W., M.C. analysed data; S.W., A.A.W.H., S.A.B., M.C. interpreted results of experiments; S.W. prepared figures and tables; S.W., A.A.W.H., S.A.B drafted manuscript; S.W., A.A.W.H., S.A.B., M.C. edited and revised manuscript.

