## Supplementary figures and images for "Blood Pressure Variability and Autonomic Response to an Acute Bout of High Intensity Interval Exercise in Healthy Young Adults"

### Supplemental Figure 1

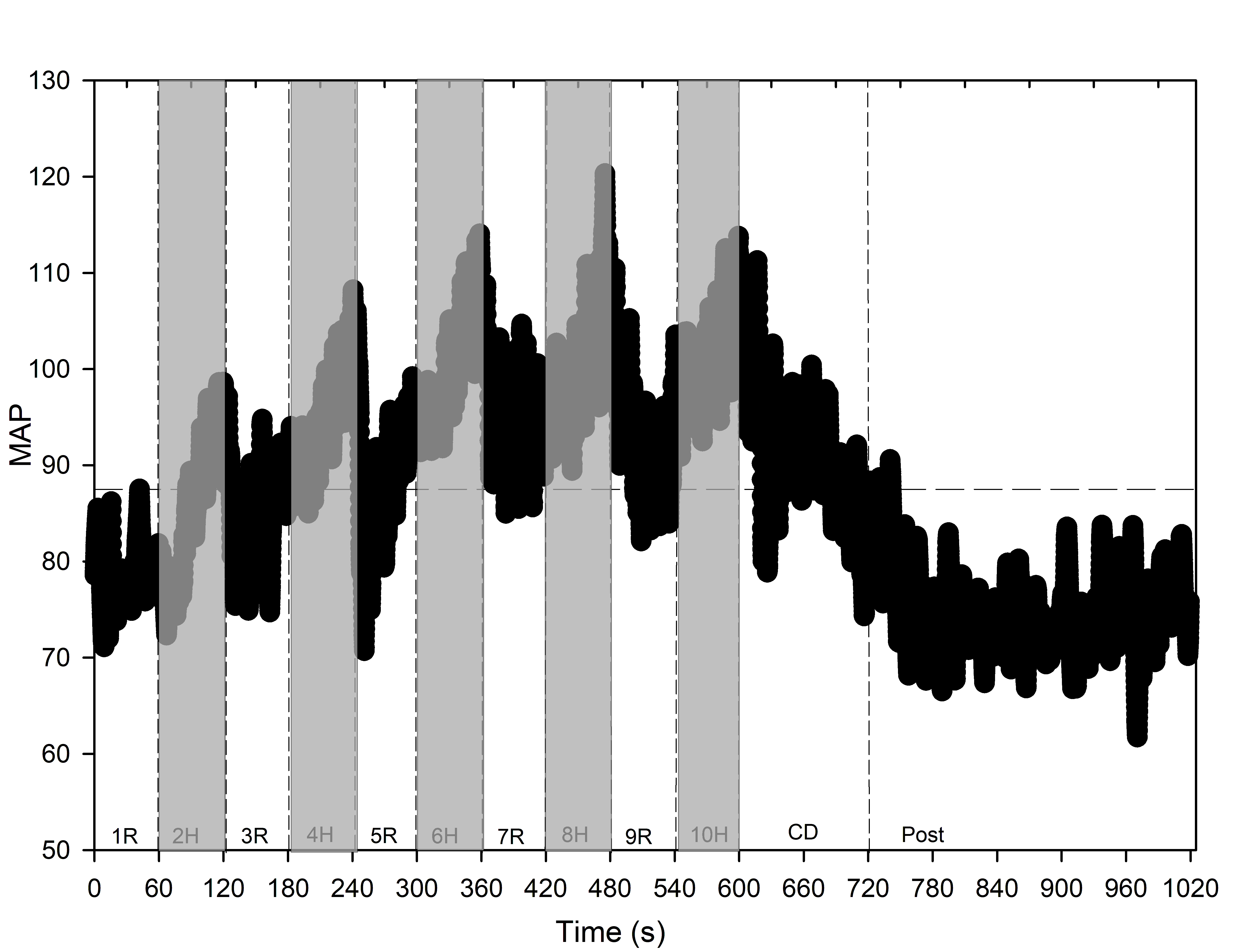

### Supplemental Figure 2

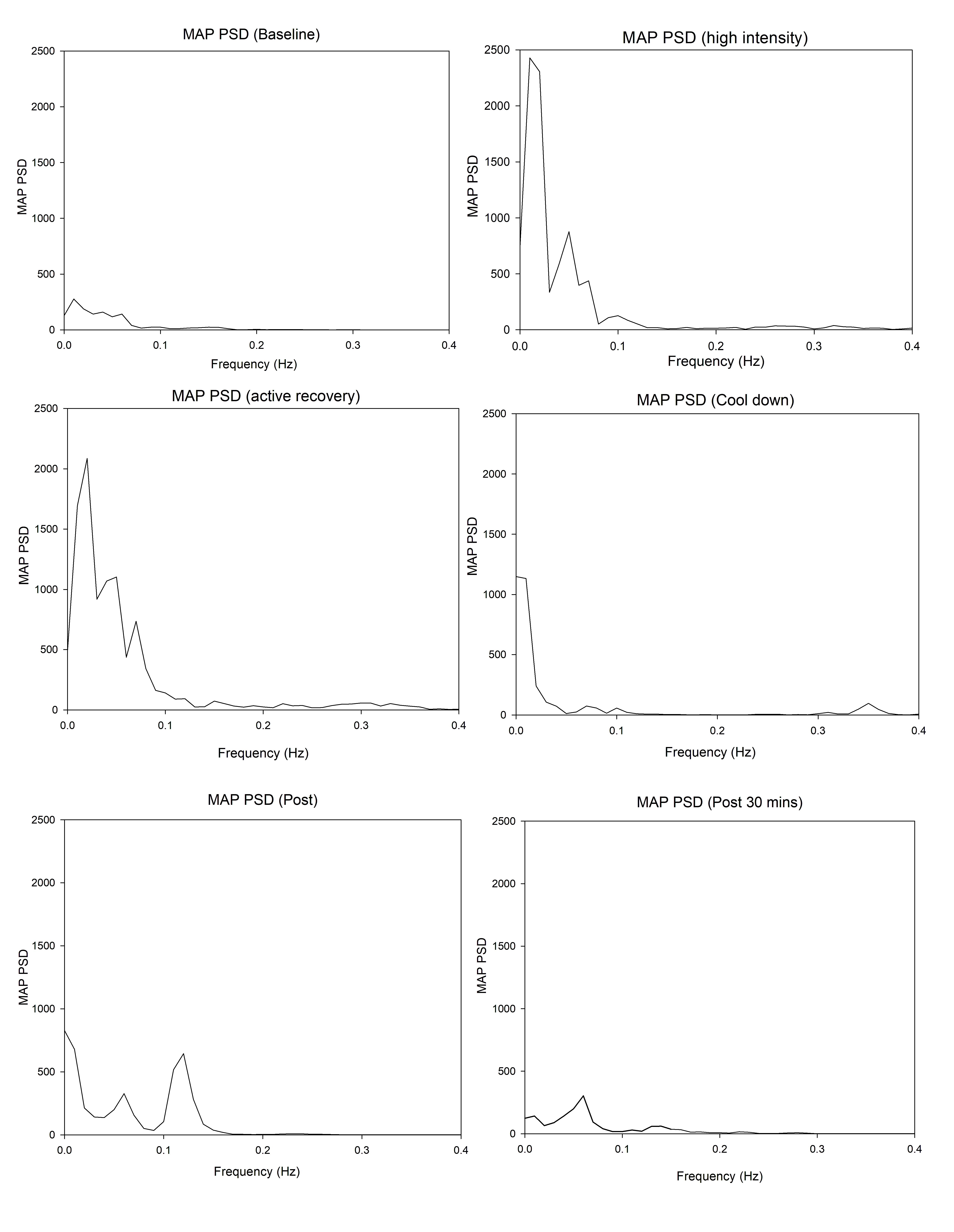
